# A Statistical Definition of Epidemic Waves

**DOI:** 10.1101/2022.05.04.22274677

**Authors:** Levente Kriston

**Affiliations:** Department of Medical Psychology, University Medical Center Hamburg-Eppendorf, Martinistr. 52, D-20246 Hamburg, Germany

**Keywords:** COVID-19, SARS-CoV-2, epidemiologic methods, Bayesian analysis, computational biology

## Abstract

The timely identification of expected surges of cases during infectious disease epidemics is essential for allocating resources and preparing interventions. This study describes a simple way to evaluate whether an epidemic wave is likely to be present based on daily new case count data. The proposed measure compares two models that assume exponential or linear dynamics, respectively. Technically, the output of two regression analyses is used to approximate a Bayes factor, which quantifies the support for the exponential over the linear model and can be used for epidemic wave detection. The trajectory of the coronavirus epidemic in three countries is analyzed and discussed for illustration. The proposed measure detects epidemic waves at an early stage, which are otherwise visible only by inspecting the development of case count data retrospectively. In addition to informing public health decision making, the outlined approach may serve as a starting point for scientific discussions on epidemic waves.

## INTRODUCTION

The course of infectious disease epidemics is frequently described by referring to ‘waves’, even though a consensual definition of what constitutes an epidemic wave is currently missing.^1–3^ Some consider the term a useful metaphor referring to a sustained upsurge (frequently called ‘spike’) in the number of sick individuals (cases).^4^ From an even broader perspective, a complete wave includes a rise in the number of cases, a defined peak, and a decline. In the present work, I focus only on the first, rising, phase of epidemic waves.

Recently, it has been suggested to use the mean of the effective reproduction number R (which refers to the average number of individuals infected by a single infectious individual during a running epidemic) over the past 14 days to operationalize epidemic waves.^3^ This working definition is certainly useful to put discussions on epidemic waves on a more objective footing. However, as the authors acknowledge, it describes rather a ‘sustained upward period’ than an upsurge in the number of cases. In addition, by calculating the average of equally weighted data points in a defined period, it discards the temporal information that is present in the data.

Technically, the unrestricted spread of infectious diseases is commonly characterized by an exponential growth of the number of confirmed cases, while reduced virus transmission and reproduction decelerates growth to a subexponential rate.^5,6^ The aim of this study was to provide a statistical measure of epidemic waves by determining whether the dynamics of an epidemic within a certain time horizon is more likely to be exponential than linear.

## METHODS

The proposed measure is based on the time series of the observed daily new cases, as using cumulative data can lead to biased conclusions.^7^ While an exponential growth of the total case counts implies an exponential growth of the daily new case counts, it is assumed here that a typical subexponential growth of the total case counts can be well approximated by a linear growth of the daily new case counts. Although the term ‘growth’ is used here, it should be noted that the suggested indicator does not differentiate between increasing and decreasing new case counts per se. Thus, it can detect not only exponential surges but also exponential decline. However, as the present study focused on epidemic surges, i.e., the increasing phase of epidemic waves, the proposed measure was calculated only if the exponent of the exponential function exceeded one (i.e., if the number of daily cases were increasing rather than declining).

The proposed epidemic wave indicator is a Bayes factor that quantifies the strength of evidence that the dynamics of an epidemic is exponential rather than linear. It is calculated using the Bayesian information criterion approximation method from the coefficient of determination of two linear models.^8^ The exponential model is calculated by regressing the logarithmic daily new case counts on time in a linear model, making use of the fact the linear association of a predictor with a logarithmic criterion is equivalent to an exponential association of the predictor and the untransformed criterion. The linear model is calculated by regressing the raw daily new case counts on time.

For the calculations, it is necessary to define a time horizon (*n* days). The wave indicator at any time point describes the development of the epidemic in the last *n* days up to the time point of calculation and should be interpreted as referring to that time interval. In the present study, the wave indicator was calculated if more than 70 percent of the daily new case numbers in the given time horizon were positive.

According to Raftery,^8^ the number of data points (*n*), the coefficient of determination (*R*^*2*^), and the number of predictors without the intercept (*p*) can be used to approximate the Bayesian information criterion (*BIC*) for linear models as

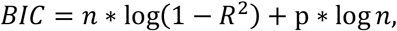

with log referring to the natural logarithm.

Following Wagenmakers,^9^ the Bayes factor (*BF*) for the support of the one model over another can be calculated from the *BIC* of the two models of as

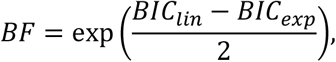

where 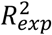 and 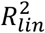 refer to the coefficient of determination in the exponential and linear models, respectively.

Merging these two equations leads to the formula

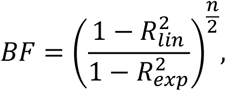

expressing the strength of support for the exponential over the linear model.

A value of one indicates that exponential and linear dynamics have the same probability. Values above one support exponential dynamics, while values below one support linear dynamics. If necessary, thresholds for interpretation are available, classifying a Bayes factor between 1 and 3 as weak, between 3 and 20 as positive, between 20 and 150 as strong, and above 150 as very strong evidence.^8^ These thresholds correspond to a 75, a 95, and a 99 percent probability that the exponential model is true, if we assume that they were equally probable before seeing the data.^9^ In the present study, a 95 percent bootstrap interval was created for the Bayes factor estimates with 500 samples in order to gain an impression on uncertainty related to the data.

All analyses were performed in R.^10^ The annotated code can be found in the Supplement.

## RESULTS

For illustration, the proposed Bayes-factor-based epidemic wave indicator was calculated for the coronavirus epidemic in the United States, in the United Kingdom, and in Germany with a time horizon of one, two, and three months, using data from the World Health Organization from initiation until February 28, 2022.^11^

Relying on daily case counts, five epidemic wave can be identified in the United States until the end of February 2022 (Figure 1). All versions of the indicator identify the first wave in March 2020 clearly. The second wave in the summer of 2020 is identified only by the 3-months version, and even that with a substantial delay in August 2020. The third wave hitting in winter 2020/21 has been clearly signaled by the 2- and 3-months versions already in November 2020. The fourth wave, which lasted through the late summer and autumn of 2021, have been recognized by all versions of the indicator around the beginning of August 2021, albeit these signals have substantial data-related uncertainty. The fifth wave of winter 2021/22 has been signaled by the 3- and (with a somewhat higher uncertainty) the 2-months versions around the turn of the year.

**Figure 1.**
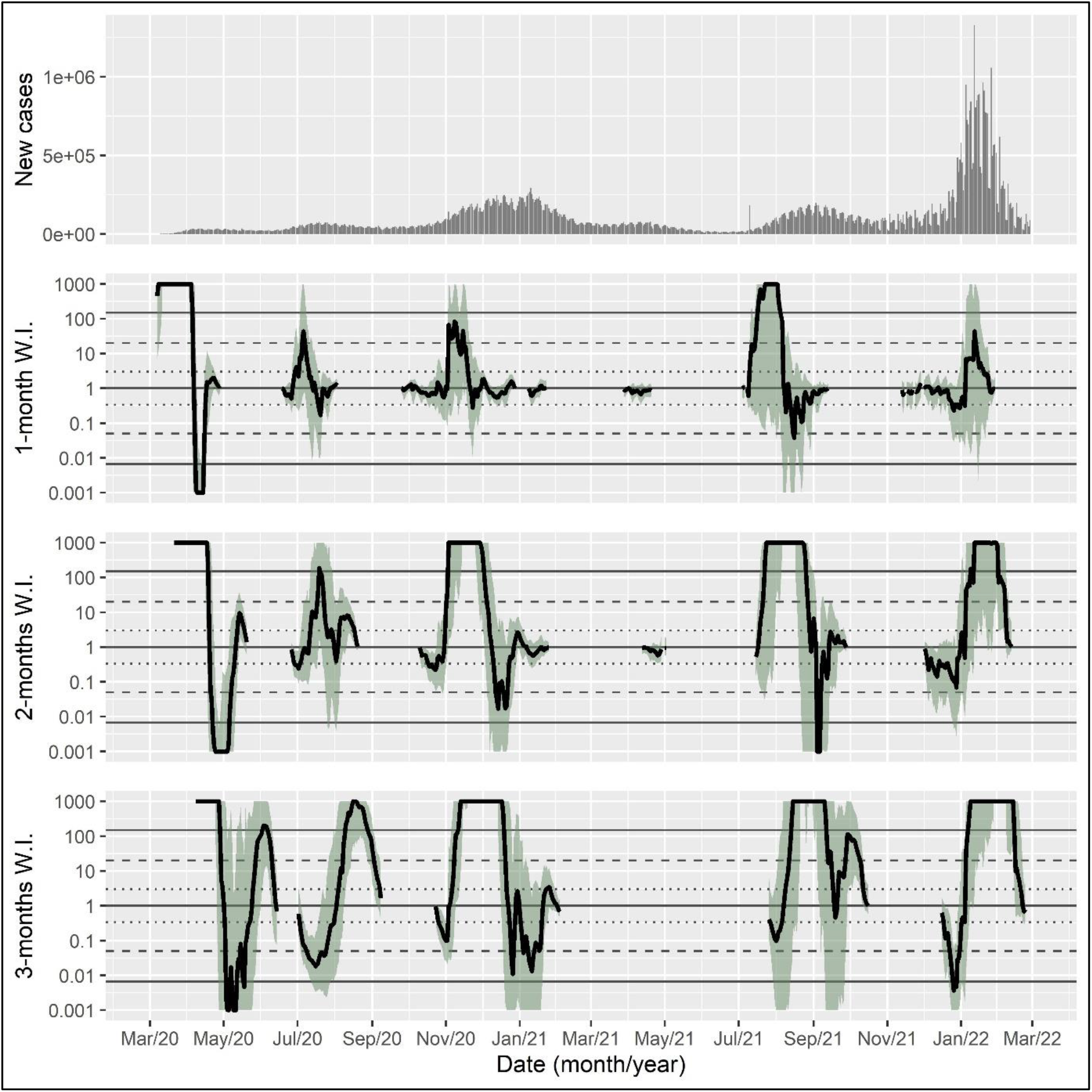
Daily new case counts and epidemic wave indicator with different time horizons in the coronavirus epidemic in the United States. *Note*. Shaded areas indicate boostrap intervals calculated with 500 samples. The dotted, dashed, and solid horizontal lines show thresholds for positive, strong, and very strong evidence, respectively. W.I, wave indicator.

Data on the daily new case counts suggest five epidemic waves in the United Kingdom up to February 2022 (Figure 2). The first surge in daily new cases in the spring of 2020 was identified as a wave irrespective of the time horizon used. The second wave in the late summer of 2020 had somewhat weaker support by the indicator. The winter wave in 2020/21 was clearly signaled by the 2- and the 3-months versions of the indicator already at the beginning of November 2020. The fourth wave on the late summer and autumn of 2021 is unequivocally recognized by all versions, with very strong evidence in July-August. The fifth wave hitting in the winter of 2021/22 is marked by very strong evidence by the 2- and 3-months versions at the beginning of January 2022.

**Figure 2.**
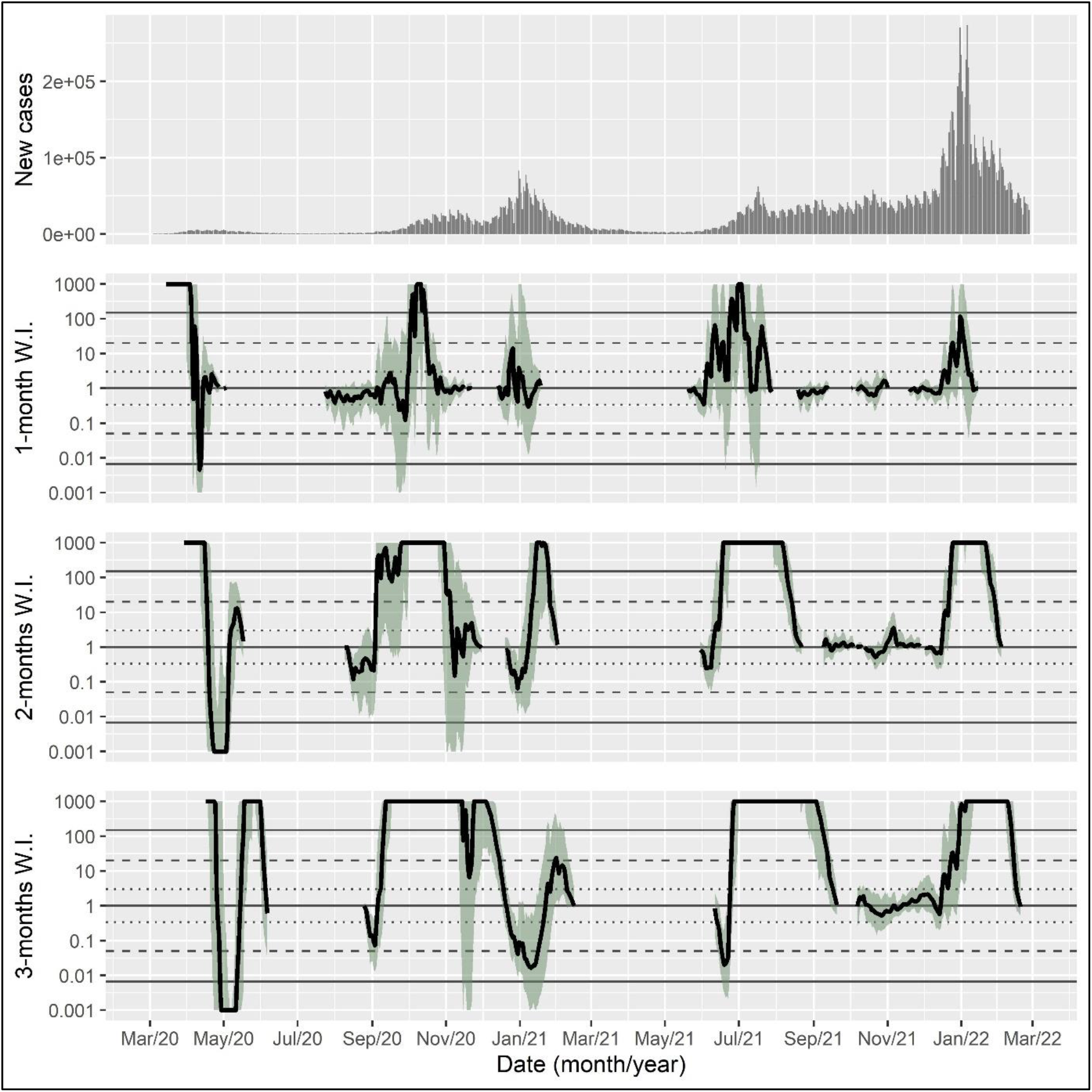
Daily new case counts and epidemic wave indicator with different time horizons in the coronavirus epidemic in the United Kingdom. *Note*. Shaded areas indicate boostrap intervals calculated with 500 samples. The dotted, dashed, and solid horizontal lines show thresholds for positive, strong, and very strong evidence, respectively. W.I, wave indicator.

The inspection of the time series of the daily new cases in Germany indicates six epidemic waves up to end of February 2022 (Figure 3). All versions of the indicator show very strong evidence of an epidemic wave in March 2020 (first wave). Very strong evidence of a second wave is provided for the autumn 2020, signaled by all versions around the middle of October 2020. A third wave that is apparent in the daily new cases data in the spring of 2021 is identified only weakly and with considerable uncertainty. A clear signal for a fourth wave is provided by the 2- and 3-months versions of the indicator in the late summer of 2021, even though the number of new cases is relatively low compared to the other waves. A fifth wave at the end of 2021 is clearly signaled only by the 3-months version, while the intensive sixth wave at the beginning of 2022 is not (yet) recognized by any version.

**Figure 3.**
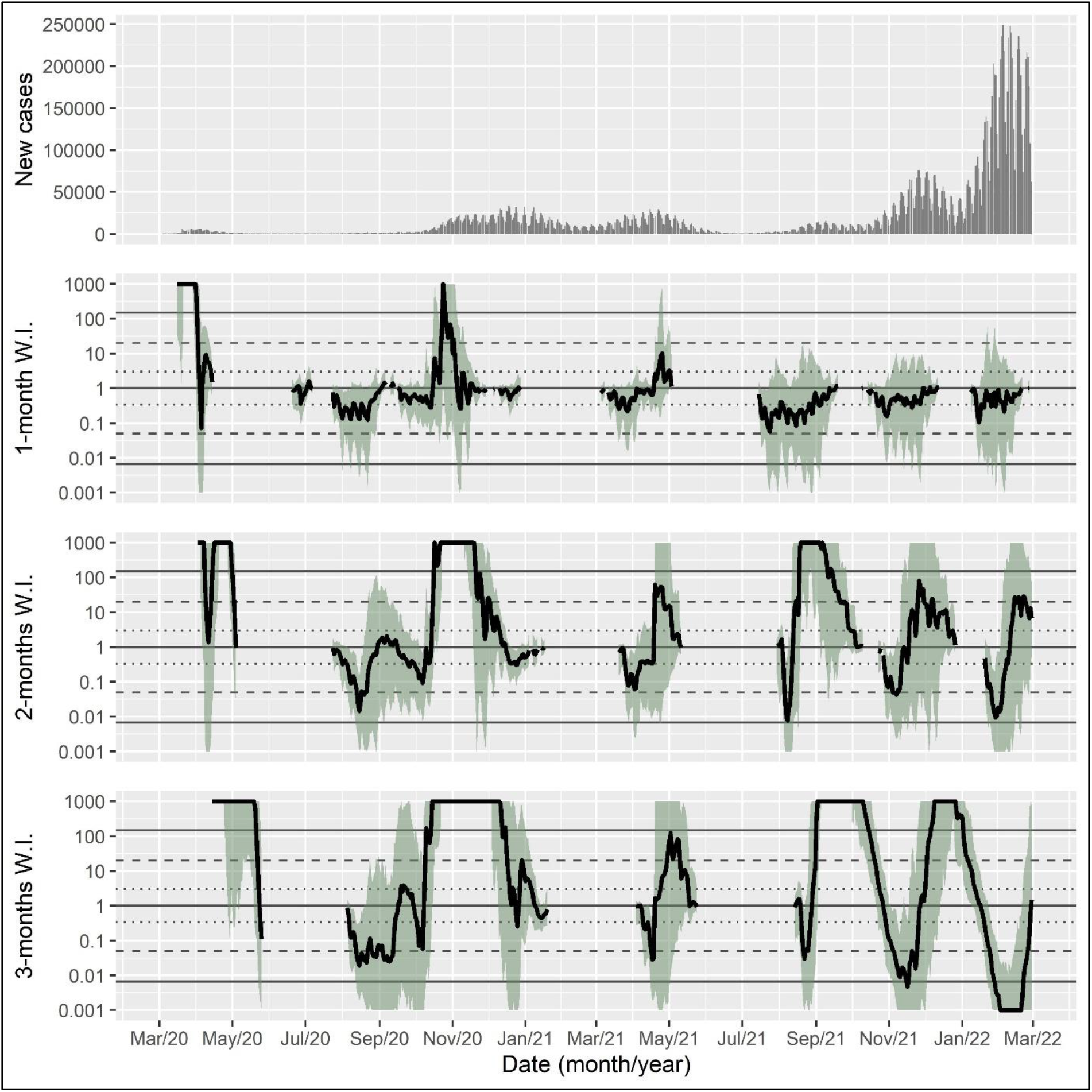
Daily new case counts and epidemic wave indicator with different time horizons in the coronavirus epidemic in Germany. *Note*. Shaded areas indicate boostrap intervals calculated with 500 samples. The dotted, dashed, and solid horizontal lines show thresholds for positive, strong, and very strong evidence, respectively. W.I, wave indicator.

Identifying epidemic waves form the time series of daily new case counts is challenging, even retrospectively. This is particularly true if the apparent waves follow each other swiftly and/or build upon each other. Instead of five to six waves as described above, data from all three countries are consistent with the interpretation of three ‘big’ waves, the first ending in the spring of 2020, the second running through autumn and winter of 2020/21, and the third centering on the winter of 2021/22. These three ‘big’ waves are all identified very clearly and early by the proposed indicator.

## DISCUSSION

The proposed approach makes clear that judgments on epidemic waves depend on the timeframe of reference and that apparently visible patterns in case count data may provide a subjective and/or incomplete picture. The measure outlined in this study is scalable to any geographic region and takes possible irregularities of the data into account.

A central limitation of the presented approach that it relies on the number of reported cases, which can be subject to inconsistencies due to variation in reporting and testing strategies. Thus, the identified waves do not necessarily reflect changes in the true number of infections. However, it is unlikely that testing and reporting strategies alone are able to produce epidemic waves with a very strong support from the proposed epidemic wave indicator. The calculation of bootstrap intervals (which should be interpreted as reference intervals rather than traditional confidence limits) can be helpful for assessing data-related uncertainty of the calculations. Still, this issue deserves further exploration.

Another challenge is posed by the question, which time horizon should be used to calculate the wave indicator. In the examples, indicators with a longer time horizon (two and three months) seem to have worked better and more clearly at detecting epidemic waves. However, the choice is likely to depend on the characteristics of the waves, of which description the measure is intended to use. For epidemics with an annual periodicity of major waves, a time horizon of several months might be appropriate. However, until clearer guidance is available, I suggest using multiple timeframes, like it was done in the present study.

An interesting characteristic of the proposed measure that it can also be used to detect phases of exponential decline in new case counts, which was not followed upon in the present study and did not have received much attention in general yet. Future modelling and empirical studies may explore whether an exponential rather than linear decline may provide valuable information regarding epidemic dynamics.

Given that even central epidemiological concepts lack a consensual definition,^12,13^ thinking about epidemic waves formally as trends with specific characteristics in time series data may be a fruitful perspective.^14^ Although the proposed measure is intended to be a descriptive indicator of epidemic waves, testing its value for prediction might be an interesting avenue of research. In addition, analyzing its agreement with similar measures, such as the average of the effective reproduction umber R across a defined period of time,^3^ could be an informative focus of future studies. Even though the presented measure is approximate, relies on simplified assumptions, and needs further evaluation, it may contribute to putting discussions on epidemic waves on a more objective basis.

## Data Availability

The dataset was derived from sources in the public domain: World Health Organization Coronavirus Disease (COVID-19) Dashboard, https://covid19.who.int.

## ADDITIONAL INFORMATION

### Author contributions

LK conceptualized and designed study, prepared the data and the software code, performed the analyses, interpreted the results, and wrote the manuscript

### Funding

No funding was received for conducting this study.

### Conflicts of interest/Competing interests

The author has no relevant financial or non-financial interests to disclose.

### Ethics approval

This is a secondary analysis of anonymized aggregate data. No ethical approval is required.

### Consent to participate

N/A

### Consent for publication

N/A

### Code availability

The custom code is attached as supplementary material.

## Notes

### Competing Interest Statement

The authors have declared no competing interest.

