## Supplement - R Code for "A Statistical Definition of Epidemic Waves"

*Levente Kriston*

```
# LIBRARIES
library(ggplot2)
library(grid)
library(caTools)
library(boot)

# OPTIONS
options(max.print=1000000)
Sys.setlocale("LC_TIME", "English")

# DATA
# Read data
data <- read.csv("https://covid19.who.int/WHO-COVID-19-global-data.csv",
                 fileEncoding = "UTF-8-BOM",
                 stringsAsFactors=F)


# Reformat datum
data$Date_reported <- as.Date(substr(data$Date_reported, 1, 10))

# BAYES FACTOR FUNCTION
# Arguments:
#   cid: country (two-letter DIN code as character)
#   n.wind: moving window width (defaults to 14)
#   n.boot: number of bootstrapping samples (defaults to 1000)
Calculate.BF <- function(cid, n.wind=14, n.boot=1000){
  # Data Sub
  data.sub <- data[which(data$Country_code==cid), ]
  # Initialize objects
  df <- list()
  coef.lin <- numeric()
  coef.exp <- numeric()
  rsq.lin <- numeric()
  rsq.exp <- numeric()
  BIC.lin <- numeric()
  BIC.exp <- numeric()
  BF <- numeric()
  bs.results <- list()
  BF.ci.ll <- numeric()
  BF.ci.ul <- numeric()
  # Loop across time points
  for (i in n.wind:nrow(data.sub)){
    # Define dataframe
    df[[i]] <- data.frame(time = c(0:(n.wind-1)),
                          raw.cases = data.sub$New_cases[(i-(n.wind-1)):i],
                          log.raw.cases = log(data.sub$New_cases[(i-(n.wind-
1)):i]))
    # Recode -Inf into missing
    df[[i]]$log.raw.cases[df[[i]]$log.raw.cases==-Inf] <- NA
    # Calculate estimates
    if (sum(df[[i]]$raw.cases==0)<(n.wind*.3)){
      coef.lin[i] <- as.numeric(lm(df[[i]]$raw.cases ~
df[[i]]$time)$coef[2])
      coef.exp[i] <- as.numeric(exp(lm(df[[i]]$log.raw.cases ~
df[[i]]$time)$coef[2]))
```

```
      rsq.lin[i] <- summary(lm(df[[i]]$raw.cases ~ df[[i]]$time))$r.squared
      rsq.exp[i] <- summary(lm(df[[i]]$log.raw.cases ~
df[[i]]$time))$r.squared
      BIC.lin[i] <- length(na.omit(df[[i]]$raw.cases))*log(1-rsq.lin[i]) +
        log(length(na.omit(df[[i]]$raw.cases)))
      BIC.exp[i] <- length(na.omit(df[[i]]$log.raw.cases))*log(1-
rsq.exp[i]) +
        log(length(na.omit(df[[i]]$log.raw.cases)))
      BF[i] <- exp((BIC.lin[i]-BIC.exp[i])/2)
    } else {
      rsq.lin[i] <- NA
      rsq.exp[i] <- NA
      BIC.lin[i] <- NA
      BIC.exp[i] <- NA
      BF[i] <- NA
    }
    # Delete values which indicate decline
    BF[coef.exp<=1] <- NA
    # Bootstrap BF confidence interval
    bs <- function(data, indices){
      d <- data[indices,]
      if (sum(d$raw.cases==0)<=(n.wind/3)){
        rsq.lin <- summary(lm(d$raw.cases ~ d$time))$r.squared
        rsq.exp <- summary(lm(d$log.raw.cases ~ d$time))$r.squared
        BIC.lin <- length(na.omit(d$raw.cases))*log(1-rsq.lin) +
          log(length(na.omit(d$raw.cases)))
        BIC.exp <- length(na.omit(d$log.raw.cases))*log(1-rsq.exp) +
          log(length(na.omit(d$log.raw.cases)))
        BF <- exp((BIC.lin-BIC.exp)/2)
        } else {
          BF <- NA
        }
      return(BF)
      }
    bs.results[[i]] <- boot(data=df[[i]],
                         statistic=bs,
                         R=n.boot)
    if (!is.na(BF[i])){
      BF.ci.ll[i] <- boot.ci(bs.results[[i]], type="perc")$perc[4]
      BF.ci.ul[i] <- boot.ci(bs.results[[i]], type="perc")$perc[5]
    } else {
      BF.ci.ll[i] <- NA
      BF.ci.ul[i] <- NA
      }
    }
    return(data.frame(date=data.sub$Date_reported,
                      cases=data.sub$New_cases,
                      coef.lin,
                      coef.exp,
                      rsq.lin,
                      rsq.exp,
                      BIC.lin,
                      BIC.exp,
                      BF,
                      BF.ci.ll,
                      BF.ci.ul)
    )
}

# PLOT FUNCTION
# Depends on the Calculate.BF function defined above
# Arguments
```

```
#   cid: country (two-letter DIN code as character)
#   n.boot: number of bootstrapping samples (defaults to 500)
#   start.date: date of beginning the x-axis
#   x.breaks: width of x-axis breaks (in ggplot nomenclature)
#   ma.wind: width of the smoothing (moving average) window
#            (defaults to 1, returns raw data without smoothing)
Plot.BF <- function(cid, n.boot=500, start.date="2020-01-01", x.breaks="2
month", ma.wind=1){
  #Calculate BFs
  BF.out.1 <- Calculate.BF(cid=cid, n.wind=28, n.boot=n.boot)
  BF.out.2 <- Calculate.BF(cid=cid, n.wind=56, n.boot=n.boot)
  BF.out.3 <- Calculate.BF(cid=cid, n.wind=84, n.boot=n.boot)
  # Subplot cases
  plot.cases <- ggplot(BF.out.1, aes(date, cases)) +
    geom_col(width=.9, fill="gray50") +
    scale_x_date(limits=c(as.Date(start.date), tail(BF.out.1$date, 1)+1),
                 breaks=x.breaks) +
    scale_y_continuous(name="New cases",
                       limits=c(0, NA)) +
    theme(
      axis.title.x = element_blank(),
      axis.text.x = element_blank(),
      axis.ticks.x=element_blank())
  # Subplot 1 month wave indicator
  BF.data.1 <- data.frame(date=BF.out.1$date,
                          BF=exp(runmean(log(BF.out.1$BF), ma.wind,
endrule="NA", align="right")),
                          BF.ci.ll=exp(runmean(log(BF.out.1$BF.ci.ll),
ma.wind, endrule="NA", align="right")),
                          BF.ci.ul=exp(runmean(log(BF.out.1$BF.ci.ul),
ma.wind, endrule="NA", align="right"))
  )
  BF.data.1$BF[BF.data.1$BF>1000] <- 1000
  BF.data.1$BF[BF.data.1$BF<1/1000] <- 1/1000
  BF.data.1$BF.ci.ul[BF.data.1$BF.ci.ul>1000] <- 1000
  BF.data.1$BF.ci.ll[BF.data.1$BF.ci.ll<1/1000] <- 1/1000
  plot.bayes.1 <- ggplot(BF.data.1, aes(date, BF))+
    geom_hline(yintercept = c(1/3, 3), linetype=3, col="gray30") +
    geom_hline(yintercept = c(1/20, 20), linetype=2, col="gray30") +
    geom_hline(yintercept = c(1/150, 1, 150), linetype=1, col="gray30") +
    geom_ribbon(aes(ymin=BF.ci.ll, ymax=BF.ci.ul),
                fill="darkseagreen4",
                alpha=.5) +
    geom_line(aes(date, BF), col="black", size=1) +
    scale_x_date(name="Date",
                 limits=c(as.Date(start.date), tail(BF.data.1$date, 1)+1),
                 breaks=x.breaks,
                 date_labels = "%b") +
    scale_y_continuous(name="1-month W.I.",
                       trans="log10",
                       breaks=10^seq(-3, 3, 1),
                       lim=c(1/(10^3), 10^3),
                       labels=c(0.001, 0.01, 0.1, 1, 10, 100, 1000))+
    theme(
      axis.title.x = element_blank(),
      axis.text.x = element_blank(),
      axis.ticks.x=element_blank())
  # Subplot 2 months wave indicator
  BF.data.2 <- data.frame(date=BF.out.2$date,
                          BF=exp(runmean(log(BF.out.2$BF), ma.wind,
endrule="NA", align="right")),
```

```
                          BF.ci.ll=exp(runmean(log(BF.out.2$BF.ci.ll),
ma.wind, endrule="NA", align="right")),
                          BF.ci.ul=exp(runmean(log(BF.out.2$BF.ci.ul),
ma.wind, endrule="NA", align="right"))
  )
  BF.data.2$BF[BF.data.2$BF>1000] <- 1000
  BF.data.2$BF[BF.data.2$BF<1/1000] <- 1/1000
  BF.data.2$BF.ci.ul[BF.data.2$BF.ci.ul>1000] <- 1000
  BF.data.2$BF.ci.ll[BF.data.2$BF.ci.ll<1/1000] <- 1/1000
  plot.bayes.2 <- ggplot(BF.data.2, aes(date, BF))+
    geom_hline(yintercept = c(1/3, 3), linetype=3, col="gray30") +
    geom_hline(yintercept = c(1/20, 20), linetype=2, col="gray30") +
    geom_hline(yintercept = c(1/150, 1, 150), linetype=1, col="gray30") +
    geom_ribbon(aes(ymin=BF.ci.ll, ymax=BF.ci.ul),
                fill="darkseagreen4",
                alpha=.5) +
    geom_line(aes(date, BF), col="black", size=1) +
    scale_x_date(name="Date",
                 limits=c(as.Date(start.date), tail(BF.data.2$date, 1)+1),
                 breaks=x.breaks,
                 date_labels = "%b") +
    scale_y_continuous(name="2-months W.I.",
                       trans="log10",
                       breaks=10^seq(-3, 3, 1),
                       lim=c(1/(10^3), 10^3),
                       labels=c(0.001, 0.01, 0.1, 1, 10, 100, 1000))+
    theme(
      axis.title.x = element_blank(),
      axis.text.x = element_blank(),
      axis.ticks.x=element_blank())
  # Subplot 3 months wave indicator
  BF.data.3 <- data.frame(date=BF.out.3$date,
                          BF=exp(runmean(log(BF.out.3$BF), ma.wind,
endrule="NA", align="right")),
                          BF.ci.ll=exp(runmean(log(BF.out.3$BF.ci.ll),
ma.wind, endrule="NA", align="right")),
                          BF.ci.ul=exp(runmean(log(BF.out.3$BF.ci.ul),
ma.wind, endrule="NA", align="right"))
  )
  BF.data.3$BF[BF.data.3$BF>1000] <- 1000
  BF.data.3$BF[BF.data.3$BF<1/1000] <- 1/1000
  BF.data.3$BF.ci.ul[BF.data.3$BF.ci.ul>1000] <- 1000
  BF.data.3$BF.ci.ll[BF.data.3$BF.ci.ll<1/1000] <- 1/1000
  plot.bayes.3 <- ggplot(BF.data.3, aes(date, BF))+
    geom_hline(yintercept = c(1/3, 3), linetype=3, col="gray30") +
    geom_hline(yintercept = c(1/20, 20), linetype=2, col="gray30") +
    geom_hline(yintercept = c(1/150, 1, 150), linetype=1, col="gray30") +
    geom_ribbon(aes(ymin=BF.ci.ll, ymax=BF.ci.ul),
                fill="darkseagreen4",
                alpha=.5) +
    geom_line(aes(date, BF), col="black", size=1) +
    scale_x_date(name="Date (month/year)",
                 limits=c(as.Date(start.date), tail(BF.data.3$date, 1)+1),
                 breaks=x.breaks,
                 date_labels = "%b/%y") +
    scale_y_continuous(name="3-months W.I.",
                       trans="log10",
                       breaks=10^seq(-3, 3, 1),
                       lim=c(1/(10^3), 10^3),
                       labels=c(0.001, 0.01, 0.1, 1, 10, 100, 1000))
  # Combine and draw
  grid.newpage()
```

```
  grid.draw(rbind(ggplotGrob(plot.cases),
                  ggplotGrob(plot.bayes.1),
                  ggplotGrob(plot.bayes.2),
                  ggplotGrob(plot.bayes.3),
                  size = "first"))
  return(list(
    plot.cases,
    plot.bayes.1,
    plot.bayes.2,
    plot.bayes.3))
}

#Example
Plot.BF("US", 200, "2020-03-01", "2 month", 1)
```
